# HPV cervical infections and serological status in vaccinated and unvaccinated women

**DOI:** 10.1101/2020.07.06.20147165

**Authors:** Carmen Lia Murall, Bastien Reyné, Christian Selinger, Claire Bernat, Vanina Boué, Sophie Grasset, Soraya Groc, Massilva Rahmoun, Noemi Bender, Marine Bonneau, Vincent Foulongne, Christelle Graf, Eric Picot, Marie-Christine Picot, Vincent Tribout, Tim Waterboer, Ignacio G Bravo, Jacques Reynes, Michel Segondy, Nathalie Boulle, Samuel Alizon

## Abstract

Understanding genital infections by Human papillomaviruses (HPVs) remains a major public health issue, especially in countries where vaccine uptake is low. We investigate HPV prevalence and antibody status in 150 women (ages 18 to 25) in Montpellier, France. At inclusion and one month later, cervical swabs, blood samples and questionnaires (for demographics and behavioural variables) were collected. Oncogenic, non-vaccine genotypes HPV51, HPV66, HPV53, and HPV52 were the most frequently detected viral genotypes overall. Vaccination status, which was well-balanced in the cohort, showed the strongest (protective) effect against HPV infections, with an associated odds ratio for alphapapillomavirus detection of 0.45 (95% confidence interval: [0.22;0.58]). We also identified significant effects of age, number of partners, body mass index, and contraception status on HPV detection and on coinfections. Type-specific IgG serological status was also largely explained by the vaccination status. IgM seropositivity was best explained by HPV detection at inclusion only. Finally, we identify a strong significant effect of vaccination on genotype prevalence, with a striking under-representation of HPV51 in vaccinated women. Variations in HPV prevalence correlate with key demographic and behavioural variables. The cross-protective effect of the vaccine against HPV51 merits further investigation.

## Introduction

Human papillomaviruses (HPVs) are the most oncogenic viruses known to infect humans [1]. This has motivated the development of safe and effective vaccines that are currently being used world-wide and target the most oncogenic genotypes (mainly HPV16 and HPV18), as well as genotypes that cause anogenital warts (HPV6 and HPV11) [2, 3]. Unfortunately, the public health burden imposed by HPVs is likely to remain high in the foreseeable future. First, in many countries, vaccine coverage is still very low (e.g. less than 25% in France in 2018 [4]). Second, as with any public health interventions against infectious diseases, it can be jeopardised by microbial diversity. In the case of HPVs, it has been argued that certain genotypes could indirectly benefit from the niches vacated following vaccination [5, 6].

Most infections by HPVs occur in young adults. However, since more than 90% of them clear within three years without having caused symptoms, they are generally considered to be benign [7, 8]. In general, we know more about the chronic and cancer stages of HPV infections than about the acute stages [9], even though the latter have implications for treating and preventing HPV-associated diseases [10]. Overall, continued surveillance of HPV genotypes and a general understanding of their natural history remain timely issues.

Within the PAPCLEAR clinical study [11], we performed an analysis of HPV infection and antibody status in women aged 18 to 25 in Montpellier, France (*N* = 149). We analysed the studied population along with three key biological variables: HPV status by PCR-based detection and genotyping at inclusion and one month later, screening for cervical lesions at inclusion, and serological status of circulating anti-HPV antibodies at inclusion. Having more than a single time point allows us to distinguish infections from transient HPV carriage. That the two visits were spread one month apart is also more informative than six or twelve months intervals, which might miss short infections [7, 12].

We analysed these data in light of vaccination status and other demographic and behavioural variables. Focusing on the effect of HPV vaccination status, we show that differential prevalence of HPVs can be detected and that changes in genotype composition are consistent with results obtained in other countries.

## Results

### Study population

Table 1 highlights key demographic characteristics of the population studied. Vaccination status is well balanced in the cohort, with 73 women (49%) being vaccinated. Among these, *n* = 63 (86%) received three doses, mostly with the tetravalent Gardasil vaccine (*n* = 62, 85%) at a median age of 15 years old. This vaccine coverage is high compared to the French national average in the corresponding cohorts, with estimates of 33.3%, 24.7%, and 5.4% for 14 years old girls born in 1993, 1994, and 1995, respectively [13]. However, most participants were students (86%) and had a high education level (40% have a BSc or a MSc), which has been reported to be associated with vaccine uptake [14].

**Table 1.** Characteristics of the *n* = 149 women included in the study stratified by vaccine status. For quantitative variables, we show the median and the 0.25 and 0.75 quantiles. One participant did not disclose her age at sexual debut. The *p*-value corresponds to differences between both groups: for continuous variables, a Wilcoxon-Mann-Whitney was used; for categorical variables we used Fisher’s exact test. *p*-values were adjusted using the Bonferroni method for multiple testing comparisons.

| Variable | | All | Vaccinated<br>$n = 73$ (49%) | unvaccinated<br>$n = 76$ (51%) | $p$-value |
| --- | --- | --- | --- | --- | --- |
| Age (years) |  | 21.7 [20.1, 23.5] | 22.0 [20.2, 23.4] | 21.3 [19.8, 23.5] | 1.00 |
| Number of partners over the last 12 months |  | 3 [2, 5] | 2 [2, 5] | 3 [2, 6] | 1.00 |
| Age at sexual debut |  | 16.0 [15.0, 17.0] | 16.0 [15.0, 17.0] | 17.0 [15.0, 17.2] | 1.00 |
| Age at menarchy (years) |  | 13.0 [12.0, 14.0] | 12.0 [12.0, 13.2] | 13.0 [12.0, 14.0] | 0.27 |
| Body Mass Index (BMI) |  | 21.2 [19.8, 23.5] | 21.9 [19.8, 24.5] | 21.0 [19.8, 22.8] | 0.93 |
| Smoking | No | $n = 95$ (64%) | $n = 48$ (66%) | $n = 47$ (62%) | 1.00 |
| | Yes | $n = 54$ (36%) | $n = 25$ (34%) | $n = 29$ (38%) | |
| Recent chlamydia infection | No | $n = 142$ (95%) | $n = 70$ (96%) | $n = 72$ (95%) | 1.00 |
| | Yes | $n = 7$ (5%) | $n = 3$ (4%) | $n = 4$ (5%) | |
| Use of a contraception method | Hormonal | $n = 90$ (60%) | $n = 42$ (58%) | $n = 48$ (63%) | 1.00 |
| | Non-hormonal | $n = 44$ (30%) | $n = 25$ (34%) | $n = 19$ (25%) | |
| | None | $n = 15$ (10%) | $n = 6$ (8%) | $n = 9$ (12%) | |

The number of new sexual partners reported over the last twelve months was high compared to other studies [15]. However, this is consistent with the longitudinal cohort inclusion criteria that participants should report at least one new sexual partner over the last year [11]. Smoking status was in line with the national average, which is 35.3% in women between 18 and 25 years [16] and so was the contraception method used, with twice as many hormonal than non-hormonal contraception users [17]. Finally, the median age at sexual debut was slightly lower than the national average, which is between 17 and 18 years [18]. The only factor that differed slightly between the vaccinated and unvaccinated populations, although non-significantly after a Bonferroni correction, was age at menarchy, which was slightly lower in the vaccinated group.

In the following, all variables listed in Table 1 are used as cofactors in the statistical tests but we only report significant effects in the main text (detailed analyses are shown in Appendix).

### Cytology

Women were screened for cervical lesions at the first visit using liquid cytology (see the Methods). In the analysis, following national recommendations, Atypical Squamous Cells of Undetermined Significance (ASCUS) were re-qualified as ‘normal’ if the sample was found to be HPV negative, and as Low-grade Squamous Intraepithelial Lesion (LSIL) if HPV positive. Twelve women (8.1%) were diagnosed with LSIL, which is consistent with French data [19], and none were found to have higher grade lesions.

The only factor associated with an increased risk of detection of a LSIL was the reported number of partners over the last twelve months (OR=1.35, 95% CI [1.02;1.81]). However, this effect could be related to our requalification of HPV positive-ASCUS as LSIL, since, as we will see, HPV detection is associated with this variable. Eight of the twelve women originally diagnosed with ASCUS were not vaccinated, but this effect was not significant (Supplementary Table S2).

### Prevalence and genotypes of HPVs

66% of the women were positive for alphapapillomavirus detection using the DEIA test in at least one of their two visits, and 47% were positive at the two visits one month apart (Table 2). This is high compared with estimates in many other countries (e.g. 24% for women less than 25 years old in [20]). However, since one of the inclusion criteria of the study was to report a new sexual partner over the last twelve months, this likely selects for a more sexually active and thus more exposed population. Vaccination status had the most significant effect on HPV detection with a strong protective effect, with an odds ratio (OR) of 0.45 and 95% confidence interval (CI) of [0.22;0.89]. The reported number of partners over the last twelve months and the participant’s age at inclusion were also associated with increased HPV detection with OR of 1.23 (95% CI [1.08;1.44]) and 1.22 (95% CI [1.02;1.47]) respectively. Counter-intuitively, participants who reported no use of contraception presented a lower odds ratio for HPV detection, although their number of new sexual partners over the last twelve months was not significantly different from that of the other participants (p-value of 0.89).

**Table 2.**
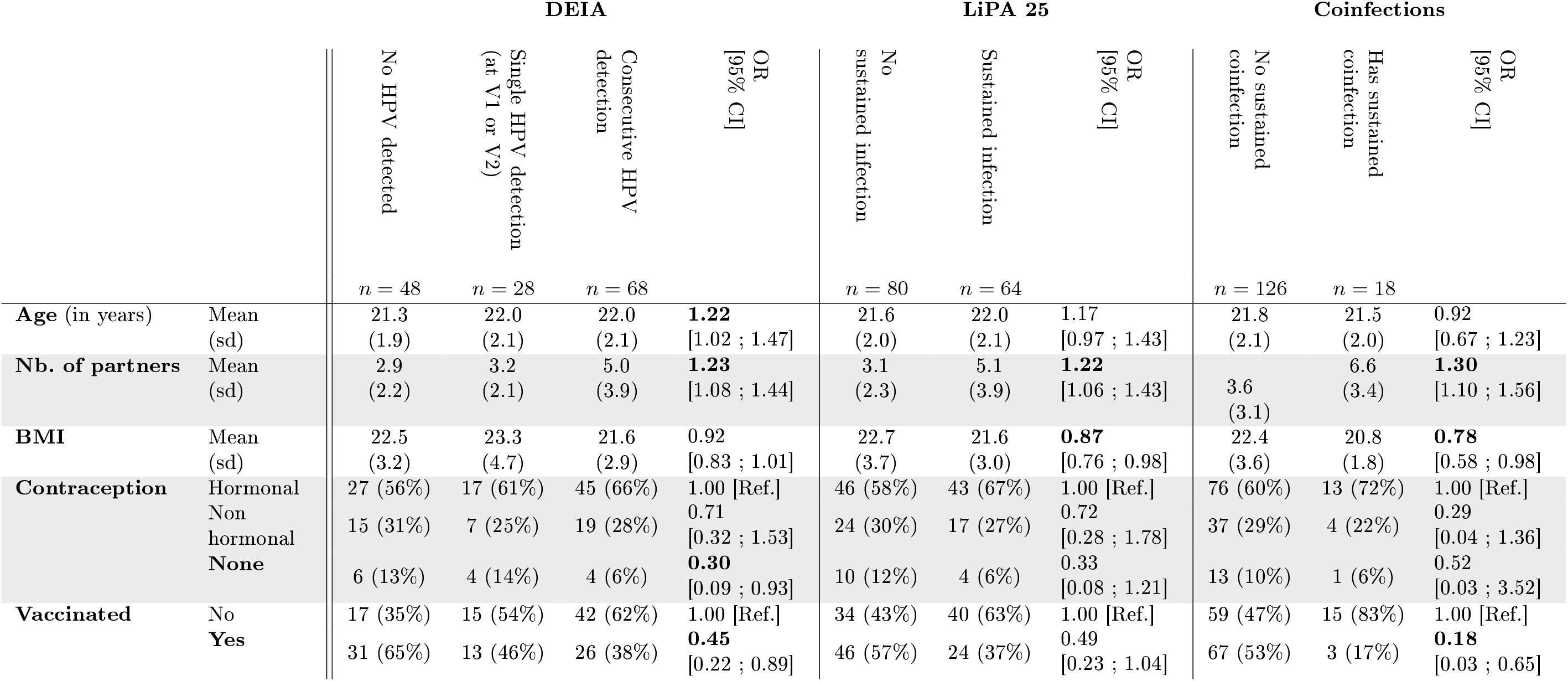
Factors associated with HPV infection. The first analysis is based on the results from the DEIA test for alphapapillomaviruses. The second analysis is based on the LiPA25 typing HPV test. The third analysis is also based on the LiPA25 results but focuses on coinfections. Overall, *n* = 144 women were included with test results at the two visits (four women were lost to follow-up after inclusion and one refused to disclose her age at sexual debut). We use ordinal logistic regression or logistic regression, depending on the number of levels of the response for the analysis. We only show here variables that are significant (in bold font) for at least one of the analyses.

The most frequent genotypes in the study population were HPV51, HPV53, and HPV66 (Figure 1). Except for the conspicuously lower prevalence of HPV16, this is consistent with the most prevalent types in France in normal cytologies being HPV16, HPV51, HPV54, and HPV53, although across all ages [20].

**Figure 1.**
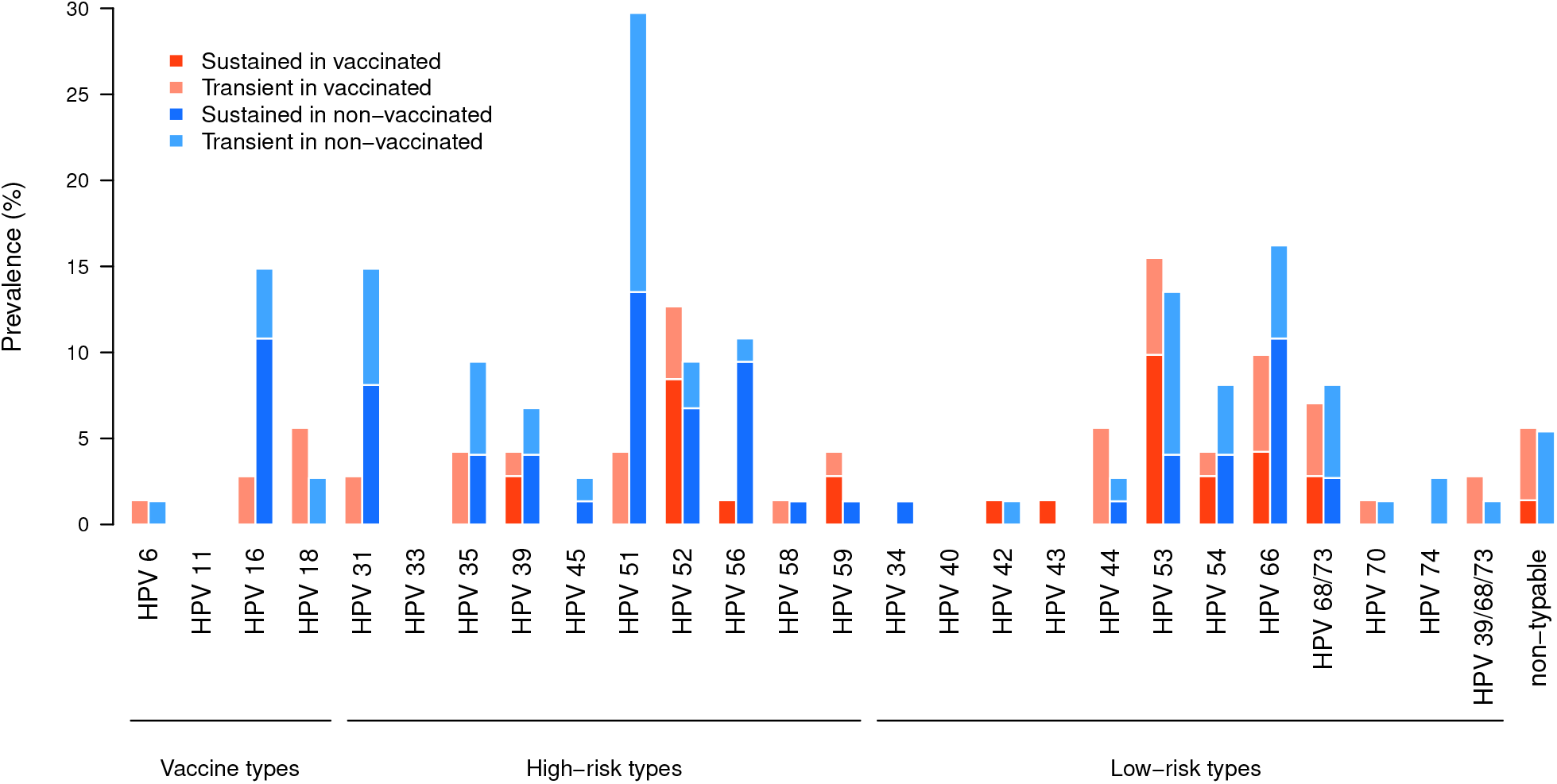
HPV genotype relative prevalence at the two visits according to vaccination status. Samples that were EIA-positive but LiPA25-negative are indicated as non-typable. We also highlight differences between genotypes that were present at both visits (dark color) or at a single visit (light color). HPVs here classified as ‘high-risk’ belong into groups 1 and 2A in the IARC carcinogenicity scheme [21].

Our testing scheme also allows to assess whether the same HPV is present in the two visits (at V1 and V2), which we refer to as ‘sustained’ infections, as opposed to ‘transient’, i.e. detected only once. As shown in Table 2, sustained infections were found in 64 women (that is 44% of the study population). This means that for most of the women who were positive twice for DEIA (94%), the repeated infection was a sustained infection. Note that for HPV18, all the infections detected were transient, whereas for HPV56 most infections were sustained. Reporting a higher number of partners over the last twelve months was associated with increased probability (OR=1.22, 95% CI [1.06;1.43]) of having a sustained infection. We also found a significant effect of Body Mass Index (BMI) towards lower probability of presenting a sustained infection (OR=0.87, 95% CI [0.76;0.98]). The effect of vaccination on presenting a sustained infection was only borderline significant with a 95% confidence interval of [0.23;1.04] for the odds ratio. However, when performing the same analysis only with oncogenic HPVs, the odds ratio for presenting a sustained infection in vaccinated women becomes 0.33 (95% CI [0.16;0.64]) (Supplementary Table S5). In this case, the odds ratio associated with the number of partners remains significant, but not the BMI effect.

Using HPV genotype information, we analysed coinfections, which we define as the detection of the same two (or more) HPV genotypes at the two visits spread one month apart. The prevalence of coinfections among all participants was 12.3%. The number of partners was positively associated with coinfections (OR=1.30, 95% CI [1.10;1.58]), while the BMI (OR=0.78, 95% CI [0.58;0.98]) and vaccination status (OR=0.18, 95% CI [0.03;0.65]) were negatively associated with coinfections (Table 2). This pattern is consistent with our results on HPV detection and sustained infections. The link with the number of partners is consistent with earlier results [15]. Note that contrary to single infections, the effect of vaccination is strongly significant without restricting the analysis to oncogenic HPVs.

### Type-specific serological status

We estimated IgM and IgG serological status specific for ten different HPVs at inclusion. As expected, IgG seropositivity was very strongly associated with vaccination status (Figure 2 and Table S8). This was true for the vaccine types but also for six other oncogenic HPVs (although to a lower extent for HPV33).

**Figure 2.**
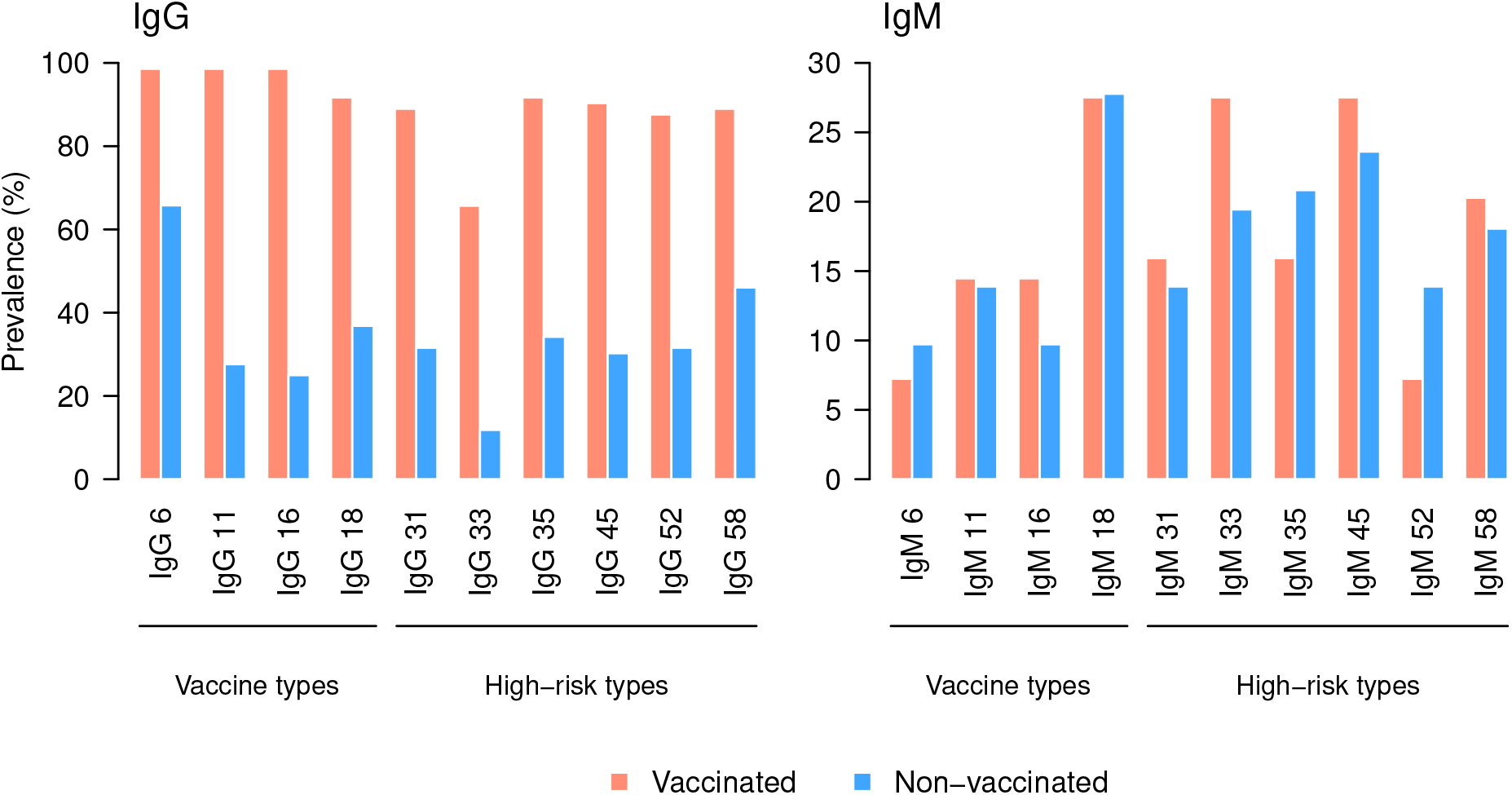
Type-specific HPV seroprevalence at inclusion. The panels show the IgG (left) and IgM (right) for vaccinated (red) and unvaccinated (blue) participants. Figures refer to the corresponding HPV, as in ‘IgG 6’ for HPV6. The antibodies detected target the L1 virus protein.

Some vaccinated participants were seronegative for the vaccine types. More precisely: one vaccinated woman presented no IgG against HPV16 and HPV18, but she only received one single dose of the vaccine at age 19; five women with a complete Gardasil vaccination schedule were seronegative for HPV18; and one woman vaccinated with Cervarix presented no immunity against HPV6 and HPV11 (which is expected given that these two types are not targeted by this vaccine). These results are consistent with the literature, especially with the lower antibody titers against HPV18 compared to those against HPV16.

To further analyse the results, we compared participants depending on the number of HPVs for which they were seropositive. For the whole population, vaccination was the only significant effect (Table S8). Focusing on unvaccinated participants, we found that participants who reported not using any contraception method were seropositive for a lower number of HPVs (Supplementary Table S9). This was consistent with these same participants having a lower risk of HPV detection (Table 2).

For IgMs, the main factor associated with being seropositive to multiple types was HPV detection at the first visit and not at the second visit one month later (Table S10). Vaccination status did not affect IgM seroprevalence, which is consistent with earlier studies [22].

### Vaccination and HPV prevalence

We found that certain HPVs were disproportionately less often detected in vaccinated women. This is the case, as expected, for vaccine-genotypes such as HPV16 (OR = 0.17, although this difference was not significant after the Bonferroni correction on testing for 25 genotypes, p-value = 0.415). We also found a similar protective effect against HPV31. Unexpectedly, the protective effect was most significant for HPV51 (OR = 0.11, p-value=0.001). The result remained significant even when we ignored transient infections and restricted the data to sustained infections. Although this result was unexpected (HPV51 is not closely related genetically to any of the vaccine types), it echoes an earlier result found on the large international PATRICIA cohort study [23].

To rule out potential issues associated with the typing test, we performed an additional PCR analysis specific for HPV51 on all the 149 inclusion visits (V1) using another protocol [24] targeting the *E7* gene instead of the *L1* gene targeted by the DEIA and LiPA_25_ assays [25]. We confirmed that vaccination status was associated with a lower prevalence of HPV51, with an odds ratio of 0.31 (95% CI [0.09,0.9], p-value = 0.023 with Fisher’s exact test).

## Discussion

HPV vaccination ignited a debate regarding the risk of so-called ‘type replacement’ [5, 26]. Indeed, since the vaccines target a limited number of genotypes, it is theoretically possible that non-vaccine genotypes may rise in prevalence occupying the niche vacated by genotypes targeted by the vaccine, either directly or indirectly due to cross-protective effects. Though technically a challenge [27, 28], it is important in the post-vaccination era that studies continue to track genotype specific prevalences [29], especially given that current vaccination programs do not cover all oncogenic HPV genotypes.

One of the difficulties in analysing the consequences of vaccination is that in most of the literature participant follow-up visits tend to be six months apart, leaving open the possibility to miss short HPV infections [7]. Here, we detected and genotyped HPVs during two visits spread one month apart. This increase in sampling density allowed us to make sure that the HPV presence was not transient, while minimising the non-detection of true, short infections. We also combined broad HPV detection and specific genotyping of 25 HPVs with the multiplex quantification of antibodies specific to ten HPVs. Unfortunately, the most prevalent HPV genotypes that we found were not present in the multiplex serological assay.

We found that broad detection of alphapapillomaviruses positively correlated with vaccination status, as well as with other expected factors, such as age or number of partners [30, 31]. HPV genotyping allowed us to refine the identification of the positive correlation with number of partners, while also revealing a negative correlation between body mass index (BMI) and the presence of HPV sustained infections. BMI has previously been reported to be associated with HPV infections, with most studies reporting either non-significant [32] or negative associations [33, 34]. Comparing different subpopulations based on key variables using Mann-Whitney (non-parametric) tests, we found no BMI difference depending on vaccination (p-value = 0.12), contraception (p-value = 0.53), and smoking (p-value = 0.48) statuses. BMI was nevertheless positively correlated with age (see Figure S1), but both factors present opposite odds ratios for HPV detection. The decreased HPV detection risk in women with higher BMI is consistent with studies that show these individuals display stronger pro-inflammatory reactions, with positive correlations between BMI and immune cell counts [35]. A meta-analysis also showed a negative association between BMI and risk of death from acute respiratory and infectious disease [36]. Here, IgM seropositivity, quantified by the number of positive type-specific responses, had a borderline-significant positive association with BMI (OR=1.11, 95% CI [0.995,1.24], Supplementary Table S10). A higher base level of immunity associated with higher BMI could provide a protection against HPV infections, which would explain the observed prevalences.

By investigating an additional marker for HPV incidence, namely seropositivity for both IgG and IgM, we showed that as expected IgG seroprevalence was largely governed by vaccination status (Table S8). For IgMs, the strongest association was with being HPV positive at the first visit only. This association with transient infections is consistent with the assumption that the infection was recent. It is more challenging to explain why IgMs would be found in participants with transient infections (positive at the first visit only) and not in sustained infections (i.e. positive at the two visits). One possibility could be that sustained infections might have been established several weeks before the inclusion visit, whereas transient infections might have been established shortly before the inclusion.

These results have direct implications for the implementation of HPV vaccination. In France, few studies have investigated HPV prevalence since the onset of vaccination, where, unfortunately, coverage remains low. More generally, the strong protective effect conferred by the vaccines against HPV51, which has already been hinted at [23], warrants further investigation, especially because this genotype is only distantly related to the vaccine types [37] and cross-protective effects were expected only in closely related types. Analysing the interaction between HPVs and the immune response (e.g. cytokines, immune cells) via longitudinal follow-up may help understand this epidemiological finding [11].

## Methods

### Data

Data originates from the PAPCLEAR study, which investigates the kinetics and ecology of HPV genital infections in young women. Here, we analyse data from the inclusion visit (Visit 1) and the result visit (Visit 2) for *N* = 149 participants (one was removed after realising she did not meet one of the inclusion criteria). The detailed protocol of the study is described elsewhere [11]. In short, during the inclusion visit, a gynaecologist or mid-wife performed a cervical smear in PerservCyt medium for squamous intraepithelial screening via liquid cytology. The same sample was used to perform HPV detection and typing (see below). Self-sample vaginal swabs and blood samples were obtained with a nurse, respectively to test for other STIs and for anti-HPV antibody assays. Women come back one month later for a results visit, where another HPV detection test is performed. Patterns of positivity at the two visits as well as population sies are shown in Figure 4.

**Figure 3.**
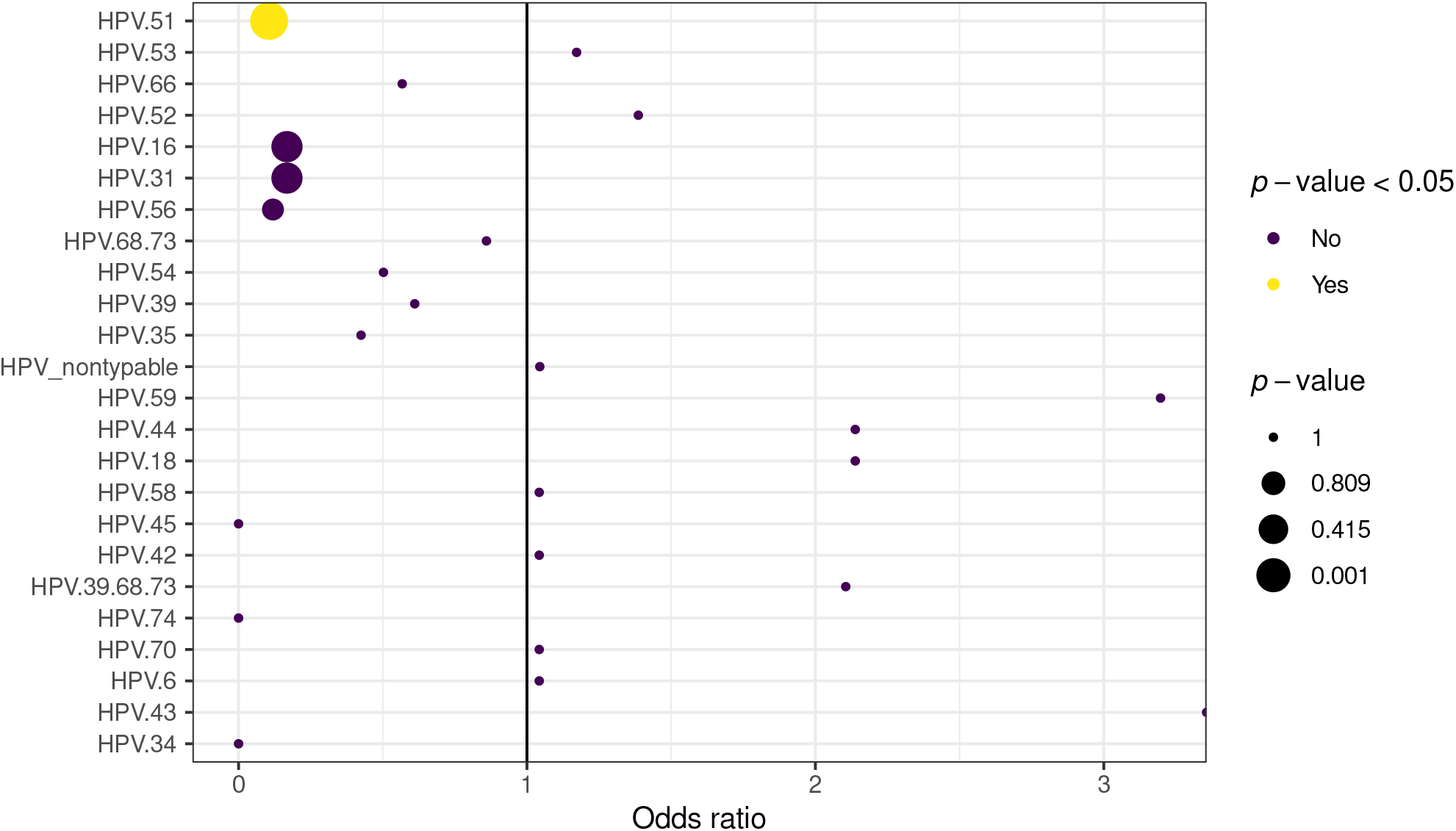
How vaccination affects the risk of detecting specific HPV genotypes. If OR=1, the HPV genotype is equally prevalent in vaccinated and unvaccinated women. If OR<1, the genotype is less prevalent in vaccinated women. Genotypes are ordered from the most often prevalent at the two visits (HPV51, on top) to the least prevalent. The size of the dot indicates the magnitude of the p-value. Significant p-values after the Bonferroni correction are in yellow.

**Figure 4:**
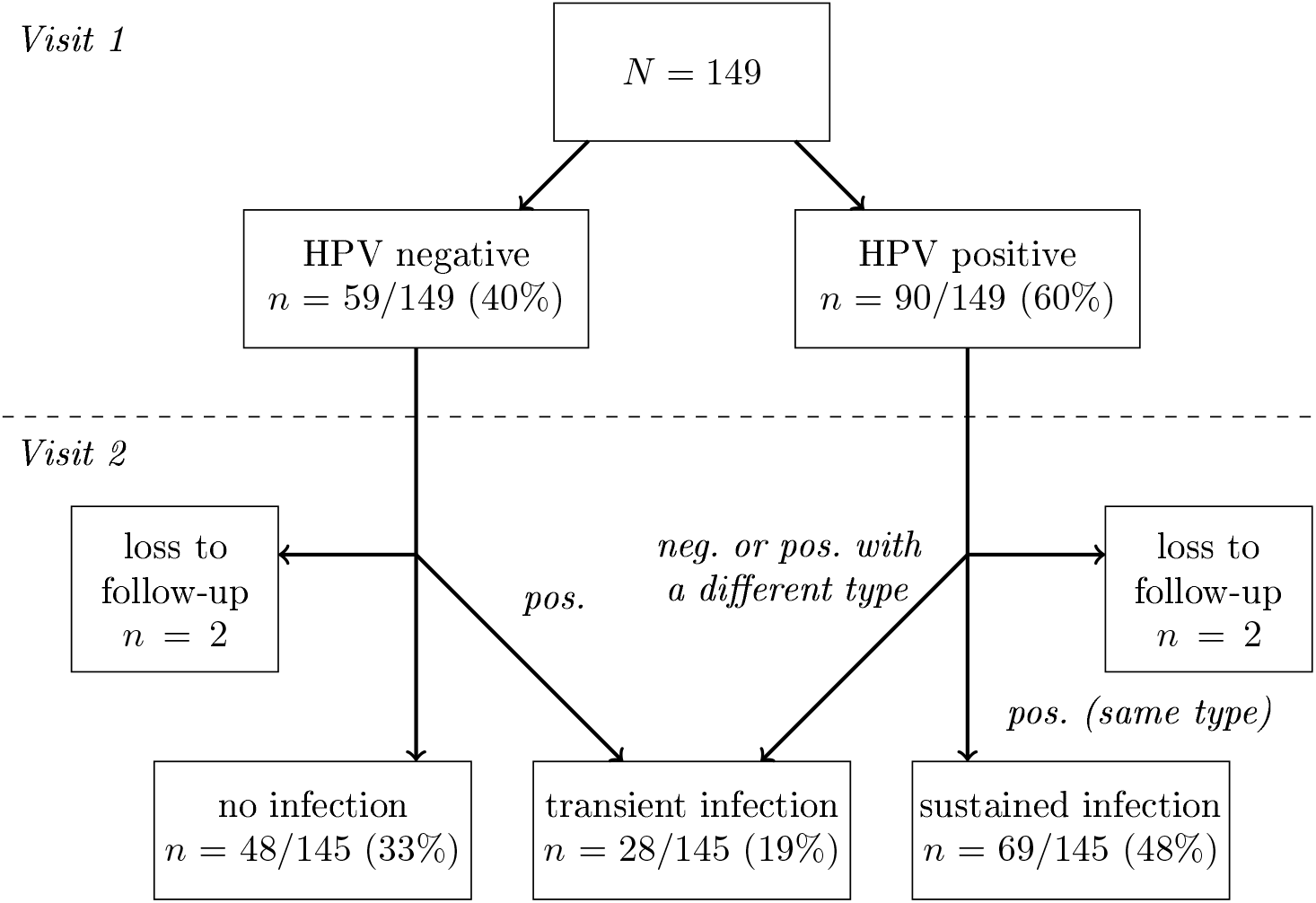
Flowchart showing the visits and the outcome of the HPV detection and typing tests.

### Protocols

For DNA extraction, we centrifuged 2mL of the cervical smear at 4000rpm for 5min. We then removed the supernatant and extracted DNA from the pellet using a QIAamp DNA Mini Kit (Qiagen51306).

For HPV detection and typing, we first tested for the presence of alphapapillomaviruses using the generic DEIA test [38]. We then used the LiPA25 typing [25] on DEIA-positive samples. Both tests are based on detecting the same amplicon of the *L1* viral gene. Total DNA in the sample was quantified using Qubit kit (Thermo Fisher Scientific). Samples that were DEIA-positive and LiPA25 negative were amplified using the PGMY PCR [39] and sequenced using Sanger sequencing. Samples for which the sequencing did not yield a clear sequence, most likely because of coinfections, are labelled as ‘non-typable’. For HPV51, we performed an additional PCR targeting the *E7* viral gene using already published primers and protocol [24].

### Protocols for circulating anti-HPV antibody

IgG and IgM antibodies against late (L1) proteins of high-risk HPV types 16, 18, 31, 33, 35, 45, 52 and 58, as well as low-risk HPV-types 6 and 11 were analysed with the multiplex serology assay using beads coated with recombinant glutathione s-transferase (GST) fusions proteins. The assay procedure has been previously described in detail [40]. Briefly, the samples were tested at a final serum dilution of 1:100 using an IgG and an IgM goat anti-Human secondary antibody. Seropositivity was defined based on standard definitions [41, 42].

### Epidemiological meta-data

Participants were asked to fill in questionnaires during the visit, and additional information was collected by the gynaecologist or mid-wife during the interview. Further details are available in the detailed protocol [11]. The variables included in this analysis are shown in Table 1.

### Statistics

We used either logistic regression or ordinal logistic regression models, depending on the number of levels of the response. For each model, we computed the odd ratios associated with each predictor along a 95% confidence interval. Note that while all variables were considered in our analyses, we only report the variables that are at least borderline significant.

For the difference in HPV genotype prevalence between the vaccinated and the unvaccinated populations, we used Fisher’s exact tests and applied a Bonferroni correction for multiple hypothesis testing.

All the analyses were performed in `R`. We used the `glm` function to conduct logistic regressions, and the `polr` function from the `MASS` package to perform ordinal logistic regressions. The proportional odds assumption was verified using the `lrm` and `residuals.lrm` functions from the `rms` package, and this assumption was verified for each of our predictors except chlamydia on the HPV presence model (which was dropped). We used the `fisher.test` function to realize Fisher’s exact tests.

### Ethics

The PAPCLEAR trial is promoted by the Centre Hospitalier Universitaire de Montpellier and has been approved by the Comitè de Protection des Personnes (CPP) Sud Mèditerranèe I on 11 May 2016 (CPP number 16 42, reference number ID RCB 2016-A00712-49); by the Comitè Consultatif sur le Traitement de l’Information en mati re de Recherche dans le domaine de la Santé on 12 July 2016 (reference number 16.504); and by the Commission Nationale Informatique et Libertès on 16 December 2016 (reference number MMS/ABD/AR1612278, decision number DR-2016-488). This trial was authorised by the Agence Nationale de Sècuritè du Mèdicament et des Produits de Santè on 20 July 2016 (reference 20160072000007). The ClinicalTrials.gov identifier is NCT02946346. All participants provided written informed consent.

## Data Availability

Data analysed in the article will be available with the published version of the article.

## Acknowledgements

The authors thank all the participants of the PAPCLEAR study and the clinical staff and nurses for their help.

## Supplementary Figure

**Figure S1.**
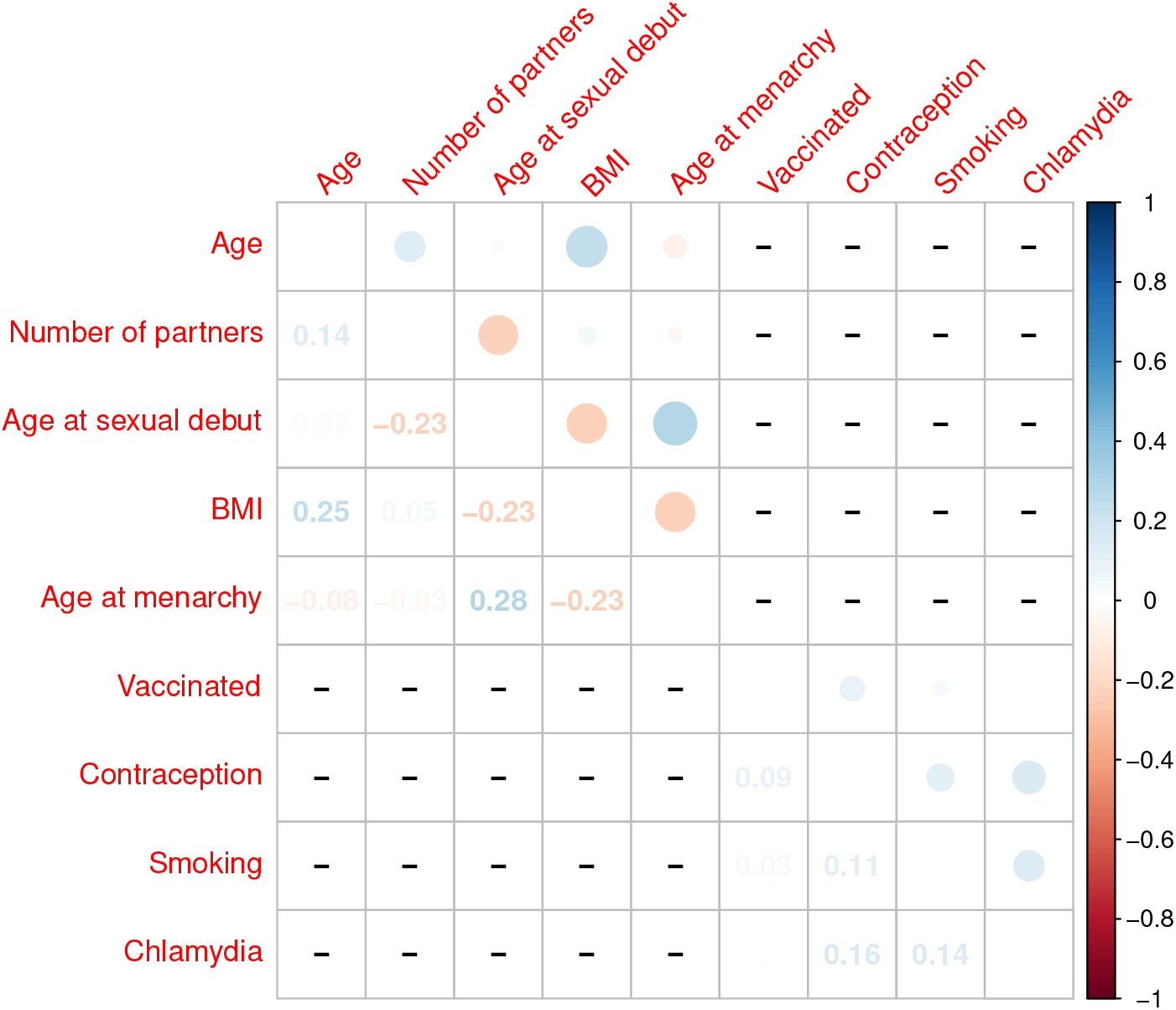
Correlations between model variables. We used Spearman’s correlation coefficient between quantitative variables and Cramèr’s V to measure the association strength between two binary/category variables. For BMI, we did not find differences based on vaccine, contraception or smoking status.

## Supplementary Tables

**Table S1.** Summary of the main variables of the statistical analyses.

| Variable |  |  | Type |
| --- | --- | --- | --- |
| Age | $q_{0.25}, q_{0.5}, q_{0.75}$ | 20.1, 21.7, 23.5 | Continuous |
| Number of partners | $q_{0.25}, q_{0.5}, q_{0.75}$ | 2, 3, 5 | Continuous |
| Age at sexual debut | $q_{0.25}, q_{0.5}, q_{0.75}$ | 15, 16, 17 | Continuous |
| Age at menarchy | $q_{0.25}, q_{0.5}, q_{0.75}$ | 12, 13, 14 | Continuous |
| BMI | $q_{0.25}, q_{0.5}, q_{0.75}$ | 19.8, 21.2, 23.5 | Continuous |
| Smoking | No | $n = 95$ | Logical |
| | Yes | $n = 54$ | |
| Chlamydia | No | $n = 140$ | Logical |
| | Yes | $n = 7$ | |
| HPV vaccination | No | $n = 76$ | Logical |
| | Yes | $n = 73$ | |
| Number of vaccine doses | 0, 1, 2, 3 |  | Discrete |
| Number of years since vaccination |  |  | Continuous |
| Contraception | No | $n = 5$ | Logical |
| | Yes | $n = 144$ | |
| HPV presence | 0 | $n = 48$ | Ordinale |
| | 1 | $n = 28$ | |
| | 2 | $n = 69$ | |
| Cytology | Normal | $n = 137$ | Categorical |
| | LSIL | $n = 12$ | |
| LiPA25 results | 0 |  | Ordinal |
|  | 1 |  |  |
|  | 2 |  |  |
| High-risk HPV | 0 | $n = 81$ | Ordinal |
| | 1 | $n = 27$ | |
| | 2 | $n = 41$ | |
| Sustained HPV infection | No | $n = 80$ | Logical |
| | Yes | $n = 65$ | |
| Sustained HPV coinfection | No | $n = 127$ | Logical |
| | Yes | $n = 18$ | |
| IgG X | No |  | Logical |
|  | Yes |  |  |
| IgM X | No |  | Logical |
|  | Yes |  |  |

**Table S2.** Factors associated with liquid cytology results.

|  |  | Normal<br>*n* = 136 | LSIL<br>*n* = 12 | OR [95% CI] |
| --- | --- | --- | --- | --- |
| Age | Mean (sd) | 21.8 (2.0) | 21.6 (2.2) | 1.04 [0.72 ; 1.49] |
| **Number of partners** | Mean (sd) | 3.9 (3.3) | 4.9 (1.9) | 1.35 [1.02 ; 1.81] |
| Age at sexual debut | Mean (sd) | 16.3 (1.8) | 15.8 (1.3) | 0.88 [0.52 ; 1.39] |
| Age at menarchy | Mean (sd) | 12.8 (1.4) | 12.8 (1.0) | 0.78 [0.39 ; 1.43] |
| BMI | Mean (sd) | 22.4 (3.4) | 21.0 (4.2) | 0.85 [0.63 ; 1.06] |
| Smoking | No | 87 (65%) | 8 (67%) | 1.00 [Ref.] |
|  | Yes | 49 (35%) | 4 (33%) | 0.79 [0.16 ; 3.38] |
| Chlamydia | No | 129 (95%) | 12 (100%) | 1.00 [Ref.] |
|  | Yes | 7 (5%) | 0 (0%) | 0 [0 ; +∞] |
| Contraception | Hormonal | 82 (60%) | 8 (67%) | 1.00 [Ref.] |
|  | Non hormonal | 43 (32%) | 0 (0%) | 0 [0 ; +∞] |
|  | None | 11 (8%) | 4 (33%) | 4.03 [0.83 ; 18.36] |
| Vaccinated | No | 68 (50%) | 8 (67%) | 1.00 [Ref.] |
|  | Yes | 68 (50%) | 4 (33%) | 0.46 [0.09 ; 1.85] |

**Table S3.** Factors associated with HPV detection. This table is based on the results of the DEIA test.

|  |  | No HPV<br>detected<br>*n* = 48 | HPV<br>detected<br>once at V1<br>or V2<br>*n* = 28 | HPV<br>detected<br>twice at V1<br>and V2<br>*n* = 68 | OR [95% CI] |
| --- | --- | --- | --- | --- | --- |
| **Age** (years) | Mean (sd) | 21.3 (1.9) | 22 (2.1) | 22 (2.1) | **1.22** [1.02 ; 1.47] |
| **Number of partners** | Mean (sd) | 2.9 (2.2) | 3.2 (2.1) | 5 (3.9) | **1.23** [1.08 ; 1.44] |
| Age at sexual debut | Mean (sd) | 16.4 (2) | 16.4 (1.7) | 16.1 (1.7) | 0.92 [0.74 ; 1.14] |
| Age at menarchy | Mean (sd) | 12.6 (1.3) | 12.8 (1.5) | 13 (1.3) | 1.11 [0.85 ; 1.45] |
| BMI | Mean (sd) | 22.5 (3.2) | 23.3 (4.7) | 21.6 (2.9) | 0.92 [0.83 ; 1.01] |
| Smoking | No | 32 (67%) | 20 (71%) | 42 (62%) | 1.00 [Ref.] |
|  | Yes | 16 (33%) | 8 (29%) | 26 (38%) | 0.75 [0.35 ; 1.57] |
| **Contraception** | Hormonal | 27 (56%) | 17 (61%) | 45 (66%) | 1.00 [Ref.] |
|  | Non hormonal | 15 (31%) | 7 (25%) | 19 (28%) | 0.71 [0.32 ; 1.53] |
|  | None | 6 (13%) | 4 (14%) | 4 (6%) | **0.3** [0.09 ; 0.93] |
| **Vaccinated** | No | 17 (35%) | 15 (54%) | 42 (62%) | 1.00 [Ref.] |
|  | Yes | 31 (65%) | 13 (46%) | 26 (38%) | **0.45** [0.22 ; 0.89] |

**Table S4.** Factors associated with HPV sustained infection. This table is based on the results of the LiPa25 HPV genotyping test.

|  |  | No sustained HPV | Has sustained HPV | OR [95% CI] |
| --- | --- | --- | --- | --- |
|  |  | *n* = 80 | *n* = 64 |  |
| Age | Mean (sd) | 21.6 (2) | 22 (2.1) | 1.17 [0.97 ; 1.43] |
| **Number of partners** | Mean (sd) | 3.1 (2.3) | 5.1 (3.9) | **1.22** [1.06 ; 1.43] |
| Age at sexual debut | Mean (sd) | 16.4 (1.8) | 16 (1.7) | 0.8 [0.61 ; 1.04] |
| Age at menarchy | Mean (sd) | 12.7 (1.4) | 13.1 (1.3) | 1.27 [0.93 ; 1.75] |
| **BMI** | Mean (sd) | 22.7 (3.7) | 21.6 (3) | **0.87** [0.76 ; 0.98] |
| Smoking | No | 54 (68%) | 40 (62%) | 1.00 [Ref.] |
|  | Yes | 26 (32%) | 24 (38%) | 0.75 [0.32 ; 1.71] |
| Chlamydia | No | 78 (98%) | 59 (92%) | 1.00 [Ref.] |
|  | Yes | 2 (2%) | 5 (8%) | 3.22 [0.51 ; 27.93] |
| Contraception | Hormonal | 46 (58%) | 43 (67%) | 1.00 [Ref.] |
|  | Non hormonal | 24 (30%) | 17 (27%) | 0.72 [0.28 ; 1.78] |
|  | None | 10 (12%) | 4 (6%) | 0.33 [0.08 ; 1.21] |
| Vaccinated | No | 34 (43%) | 40 (63%) | 1.00 [Ref.] |
|  | Yes | 46 (57%) | 24 (37%) | 0.49 [0.23 ; 1.04] |

**Table S5.** Factors associated with sustained infection by high-risk HPVs. This table is based on the results of the LiPa25 HPV genotyping test.

|  |  | No high-risk HPV detected | High-risk HPV detected once | Same high-risk HPV detected twice | OR [95% CI] |
| --- | --- | --- | --- | --- | --- |
|  |  | *n* = 76 | *n* = 27 | *n* = 41 |  |
| Age | Mean (sd) | 21.6 (2.0) | 22.2 (2.3) | 21.9 (2.0) | 1.12 [0.95 ; 1.33] |
| **Number of partners** | Mean (sd) | 3.5 (3.2) | 3.5 (3.2) | 5.1 (3.3) | **1.12** [1.003 ; 1.27] |
| Age at sexual debut | Mean (sd) | 16.4 (1.9) | 16.2 (1.7) | 16.1 (1.6) | 0.92 [0.74 ; 1.13] |
| Age at menarchy | Mean (sd) | 12.8 (1.3) | 12.6 (1.5) | 13.2 (1.2) | 1.07 [0.82 ; 1.41] |
| BMI | Mean (sd) | 22.1 (3.3) | 23.1 (4.3) | 21.8 (3.0) | 1 [0.9 ; 1.1] |
| Smoking | No | 46 (61%) | 21 (78%) | 27 (66%) | 1.00 [Ref.] |
|  | Yes | 30 (39%) | 6 (22%) | 14 (34%) | 0.5 [0.23 ; 1.04] |
| Chlamydia | No | 72 (95%) | 26 (96%) | 39 (95%) | 1.00 [Ref.] |
|  | Yes | 4 (5%) | 1 (4%) | 2 (5%) | 0.69 [0.12 ; 3.49] |
| Contraception | Hormonal | 46 (60%) | 15 (56%) | 28 (69%) | 1.00 [Ref.] |
|  | Non hormonal | 22 (29%) | 9 (33%) | 10 (24%) | 0.72 [0.32 ; 1.6] |
|  | None | 8 (11%) | 3 (11%) | 3 (7%) | 0.52 [0.15 ; 1.66] |
| **Vaccinated** | No | 30 (39%) | 14 (52%) | 30 (73%) | 1.00 [Ref.] |
|  | **Yes** | 46 (61%) | 13 (48%) | 11 (27%) | **0.33** [0.16 ; 0.64] |

**Table S6.** Factors associated with sustained HPV coinfection.

|  |  | No sustained<br>coinfection<br>*n* = 126 | Has sustained<br>coinfection<br>*n* = 18 | OR [95% CI] |
| --- | --- | --- | --- | --- |
| Age | Mean (sd) | 21.8 (2.1) | 21.5 (2.0) | 0.92 [0.67 ; 1.23] |
| **Number of partners** | Mean (sd) | 3.6 (3.1) | 6.6 (3.4) | **1.30** 1.3 [1.1 ; 1.56] |
| Age at sexual debut | Mean (sd) | 16.3 (1.7) | 15.9 (2.0) | 0.88 [0.61 ; 1.25] |
| Age at menarchy | Mean (sd) | 12.8 (1.3) | 13.1 (1.5) | 0.98 [0.57 ; 1.63] |
| **BMI** | Mean (sd) | 22.4 (3.6) | 20.8 (1.8) | **0.78** [0.58 ; 0.98] |
| Smoking | No | 85 (67%) | 9 (50%) | 1.00 [Ref.] |
|  | Yes | 41 (33%) | 9 (50%) | 1.84 [0.55 ; 6.28] |
| Chlamydia | No | 121 (96%) | 16 (89%) | 1.00 [Ref.] |
|  | Yes | 5 (4%) | 2 (11%) | 2.2 [0.18 ; 23.75] |
| Contraception | Hormonal | 76 (60%) | 13 (72%) | 1.00 [Ref.] |
|  | Non hormonal | 37 (29%) | 4 (22%) | 0.29 [0.04 ; 1.36] |
|  | None | 13 (10%) | 1 (6%) | 0.52 [0.03 ; 3.52] |
| **Vaccinated** | No | 59 (47%) | 15 (83%) | 1.00 [Ref.] |
|  | Yes | 67 (53%) | 3 (17%) | **0.18**[0.03 ; 0.65] |

**Table S7.**
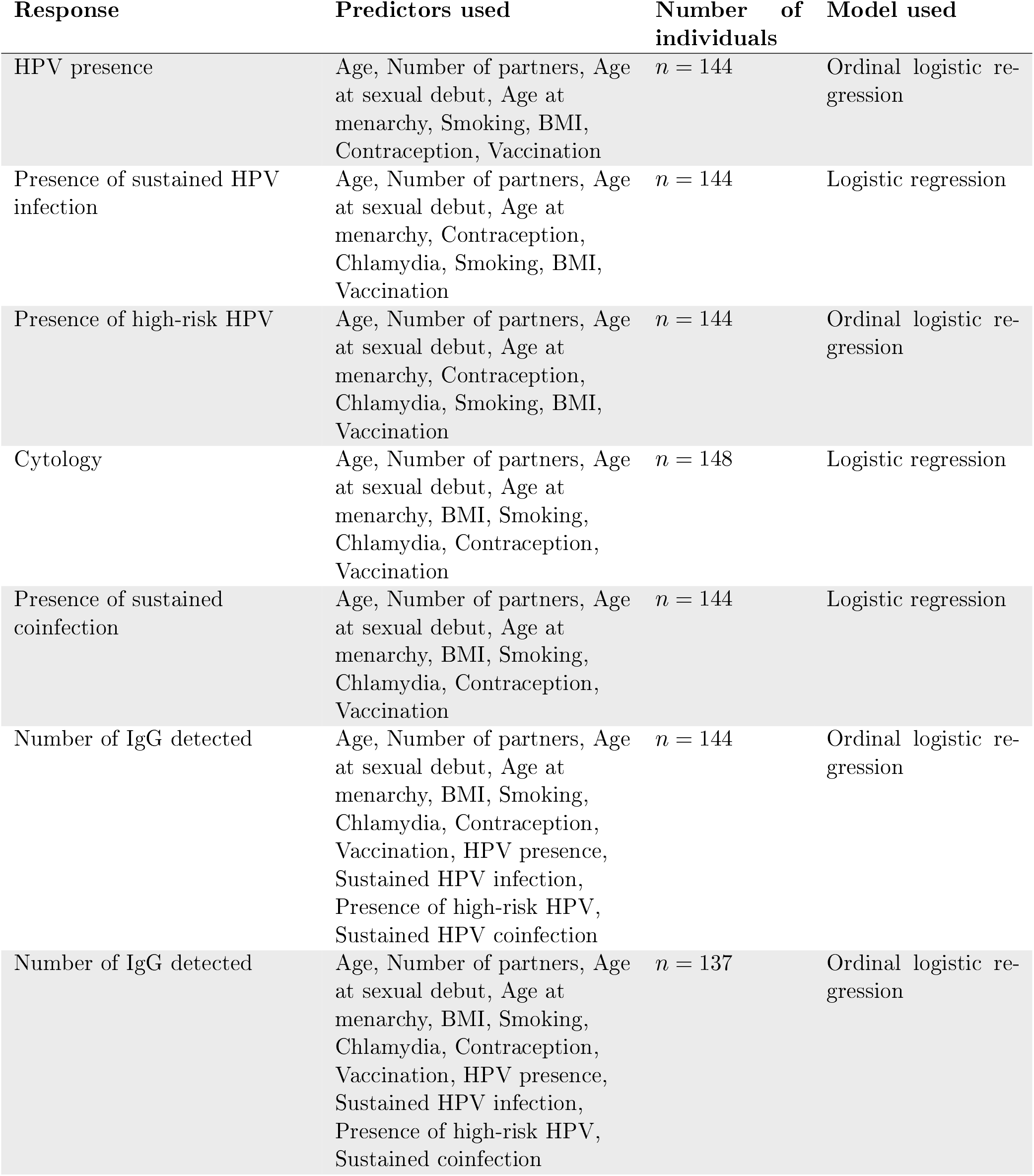
Summary of the statistical analyses performed.

**Table S8.**
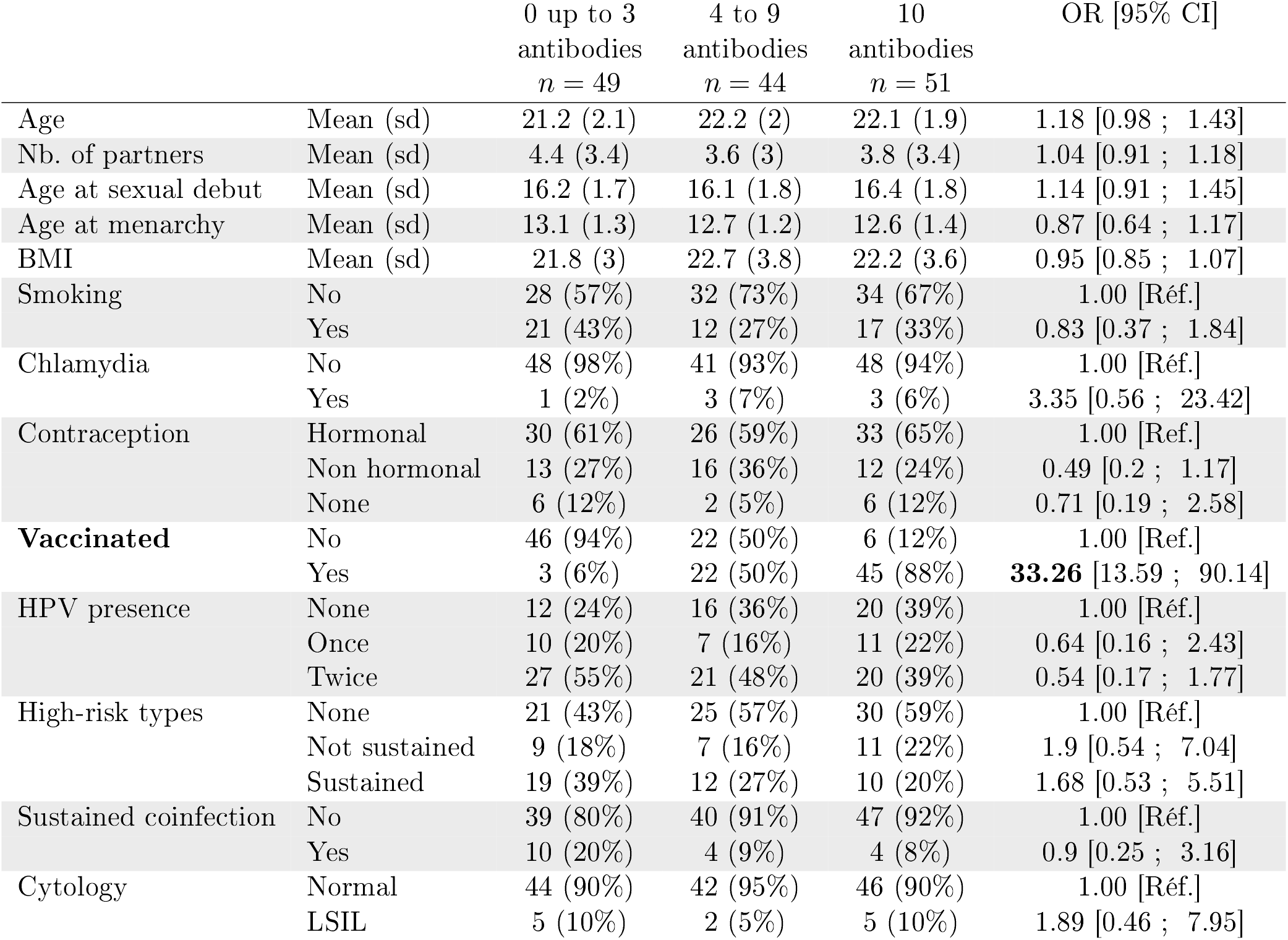
Number of IgG above positivity threshold among all participants.

**Table S9.** Number of IgG antibodies above positivity threshold among unvaccinated women.

|  |  | 0 or 1<br>antibody<br>*n* = 19 | 2 or 3<br>antibodies<br>*n* = 27 | 4 up to 10<br>antibodies<br>*n* = 28 | OR [95% CI] |
| --- | --- | --- | --- | --- | --- |
| Age | Mean (sd) | 21.1 (2) | 21.3 (2.2) | 22.3 (2) | 1.23 [0.98 ; 1.55] |
| Nb. of partners | Mean (sd) | 4.6 (4) | 4.4 (3.2) | 4.1 (3.2) | 0.95 [0.79 ; 1.13] |
| Age at sexual debut | Mean (sd) | 16.4 (2.1) | 16 (1.4) | 16.5 (1.9) | 1.08 [0.79 ; 1.49] |
| Age at menarchy | Mean (sd) | 13.1 (1.3) | 13.3 (1.4) | 12.9 (1) | 0.84 [0.57 ; 1.22] |
| BMI | Mean (sd) | 21.9 (3.4) | 21.4 (2.6) | 22.2 (4.1) | 1.03 [0.89 ; 1.21] |
| Smoking | No | 11 (58%) | 16 (59%) | 19 (68%) | 1.00 [Réf.] |
|  | Yes | 8 (42%) | 11 (41%) | 9 (32%) | 0.74 [0.27 ; 2] |
| Chlamydia | No | 18 (95%) | 27(100%) | 25 (89%) | 1.00 [Réf.] |
|  | Yes | 1 (5%) | 0 (0%) | 3 (11%) | 4.51 [0.41 ; 115.3] |
| **Contraception** | Hormonal | 10 (53%) | 17 (63%) | 21 (75%) | 1.00 [Ref.] |
|  | Non hormonal | 5 (26%) | 8 (30%) | 5 (18%) | 0.84 [0.26 ; 2.64] |
|  | **None** | 4 (21%) | 2 (7%) | 2 (7%) | **0.15** [0.02 ; 0.83] |
| HPV presence | None | 8 (42%) | 3 (11%) | 6 (21%) | 1.00 [Réf.] |
|  | Once | 3 (16%) | 6 (22%) | 6 (21%) | 2.38 [0.41 ; 14.33] |
|  | Twice | 8 (42%) | 18 (67%) | 16 (57%) | 0.9 [0.17 ; 4.95] |
| High-risk types | None | 12 (63%) | 8 (30%) | 10 (36%) | 1.00 [Réf.] |
|  | Not sustained | 3 (16%) | 5 (19%) | 6 (21%) | 0.9 [0.18 ; 4.34] |
|  | Sustained | 4 (21%) | 14 (52%) | 12 (43%) | 2.38 [0.55 ; 10.66] |
| Sustained coinfection | No | 16 (84%) | 20 (74%) | 23 (82%) | 1.00 [Réf.] |
|  | Yes | 3 (16%) | 7 (26%) | 5 (18%) | 0.67 [0.16 ; 2.89] |
| Cytology | Normal | 18 (95%) | 23 (85%) | 25 (89%) | 1.00 [Réf.] |
|  | LSIL | 1 (5%) | 4 (15%) | 3 (11%) | 1.63 [0.36 ; 7.66] |

**Table S10.** Number of IgM antibodies above positivity threshold in all participants.

|  |  | 0<br>antibodies<br>*n* = 69 | 1 or 2<br>antibodies<br>*n* = 35 | 3 up to 10<br>antibodies<br>*n* = 32 | OR [95% CI] |
| --- | --- | --- | --- | --- | --- |
| Age | Mean (sd) | 21.9 (2) | 21.7 (2.2) | 21.6 (2.2) | 0.87 [0.73 ; 1.04] |
| Number of partners | Mean (sd) | 3.9 (3.2) | 4.2 (3.7) | 4.3 (3.1) | 1.03 [0.92 ; 1.15] |
| Age at sexual debut | Mean (sd) | 16.3 (1.8) | 16.4 (1.8) | 15.9 (1.4) | 0.95 [0.76 ; 1.17] |
| Age at menarchy | Mean (sd) | 12.6 (1.2) | 12.9 (1.3) | 12.9 (1.6) | 1.19 [0.89 ; 1.58] |
| BMI | Mean (sd) | 21.9 (3.2) | 21.7 (2.7) | 23.3 (4.5) | 1.11 [0.995 ; 1.24] |
| Smoking | No | 44 (64%) | 22 (63%) | 20 (62%) | 1.00 [Ref.] |
|  | Yes | 25 (36%) | 13 (37%) | 12 (38%) | 0.97 [0.46 ; 2.04] |
| Chlamydia | No | 68 (99%) | 32 (91%) | 29 (91%) | 1.00 [Ref.] |
|  | Yes | 1 (1%) | 3 (9%) | 3 (9%) | 2.6 [0.57 ; 13.02] |
| Contraception | Hormonal | 40 (58%) | 21 (60%) | 22 (69%) | 1.00 [Ref.] |
|  | Non hormonal | 23 (33%) | 10 (29%) | 7 (22%) | 0.68 [0.3 ; 1.54] |
|  | None | 6 (9%) | 4 (11%) | 3 (9%) | 0.97 [0.3 ; 2.98] |
| Vaccinated | No | 39 (57%) | 16 (46%) | 17 (53%) | 1.00 [Ref.] |
|  | Yes | 30 (43%) | 19 (54%) | 15 (47%) | 1.45 [0.71 ; 3.01] |
| **HPV presence** | Not V1 nor V2 | 25 (36%) | 14 (40%) | 5 (16%) | 1.00 [Ref.] |
|  | **Just V1** | 7 (10%) | 2 (6%) | 9 (28%) | **4.9** [1.29 ; 19.94] |
|  | Just V2 | 4 (6%) | 2 (6%) | 2 (6%) | 2.87 [0.43 ; 18.53] |
|  | Twice | 33 (48%) | 17 (49%) | 16 (50%) | 2.43 [0.79 ; 7.5] |
| High-risk HPV | None | 36 (52%) | 18 (51%) | 17 (53%) | 1.00 [Ref.] |
|  | Not sustained | 13 (19%) | 4 (11%) | 8 (25%) | 0.53 [0.15 ; 1.8] |
|  | Sustained | 20 (29%) | 13 (37%) | 7 (22%) | 0.58 [0.19 ; 1.78] |
| Sustained coinfection | No | 60 (87%) | 29 (83%) | 29 (91%) | 1.00 [Ref.] |
|  | Yes | 9 (13%) | 6 (17%) | 3 (9%) | 0.78 [0.24 ; 2.42] |

